# Durability of response to the third dose of the SARS-CoV-2 BNT162b2 vaccine in adults aged 60 years and older: Three-month follow-up

**DOI:** 10.1101/2021.12.25.21268336

**Authors:** Noa Eliakim-Raz, Amos Stemmer, Yaara Leibovici-Weisman, Asaf Ness, Muhammad Awwad, Nassem Ghantous, Noam Erez, Avital Bareket-Samish, Adva Levy-Barda, Haim Ben-Zvi, Neta Moskovits, Erez Bar-Haim, Salomon M. Stemmer

## Abstract

**Background:** Age/frailty are strong predictors of COVID-19 mortality. After the second BNT162b2 dose, immunity wanes faster in older (≥65 years) versus younger adults. The durability of response after the third vaccine is unclear.

**Methods:** This prospective cohort study included healthcare workers/family members≥60 years who received a third BNT162b2 dose. Blood samples were drawn immediately before (T0), 10□19 (T1), and 74□103 (T2) days after the third dose. Anti-spike IgG titers were determined using a commercial assay, seropositivity was defined as≥50 AU/mL. Neutralising antibody titres were determined at T2. Adverse events, COVID-19 infections, and clinical frailty scale (CFS) levels were documented.

**Findings:** The analysis included 97 participants (median age, 70 years [IQR, 66□74], 58% CFS level 2). IgG titres, which increased significantly from T0 to T1 (medians, 440 AU/mL [IQR, 294□923] and 25,429 [14203□36114] AU/mL, respectively; p<0·001), decreased significantly by T2, but all remained seropositive (median, 8306 AU/mL [IQR, 4595□14701], p<0·001 *vs* T1). In a multivariable analysis, only time from the first vaccine was significantly associated with lower IgG levels at T2 (p=0·004). At T2, 60 patients were evaluated for neutralising antibodies; all were seropositive (median, 1294 antibody titre [IQR, 848□2072]). Neutralising antibody and anti-spike IgG levels were correlated (R=0·6, p<0·001). No major adverse events or COVID-19 infections were reported.

**Interpretation:** Anti-spike IgG and neutralising antibody levels remain adequate 3 months after the third BNT162b2 vaccine in healthy adults≥60 years, although the decline in IgG is concerning. A third vaccine dose in this population should be top priority.

**Funding:** No external funding.

**Research in context:** *Evidence before this study:* We searched PubMed on Aug 1, 2021, for published research articles with no date restrictions, using the search terms of “SARS-Cov-2”, “COVID-19”, “vaccine”, “dose”, “antibody response”, and “adults” with English as a filter. Several studies were identified that investigated waning of immunity in healthy adults. It is well established through epidemiology and serology studies that in healthy adults, the protection conferred by the BNT162b2 messenger RNA (mRNA) vaccine (Pfizer/BioNtech) wanes significantly after several months. Studies have also shown that the immune response to the vaccine varies with age, and that after the second dose of the BNT162b2 vaccine, the older adult population (65-85 years of age) typically has a lower immune response (as reflected in an analysis of anti-spike IgG antibodies and neutralising antibody titres), than younger adults (18-55 years of age), and that the immunity wanes in all age groups within several months.

*Added value of this study:* This is, to our knowledge, the first study that examined anti-spike IgG and neutralising antibody titres three months after the third BNT162b2 vaccine dose. The study has demonstrated that three months after that dose, participants, who were healthy adults aged 60 years and older, remained anti-spike IgG seropositive, although a significant decrease in anti-spike IgG titres was observed (compared to two weeks after the third dose). In addition, a statistically significant correlation was observed between the neutralising antibody titres and the anti-spike IgG titres, and all participants were seropositive for neutralising antibodies three months after the third dose. Also, no major adverse events or COVID-19 infections were reported.

*Implications of all the available evidence:* As our data suggest that a third dose of the BNT162b2 vaccine is effective in maintaining adequate immune response against COVID-19 for at least several months in healthy adults aged 60 years and older, and as it is well established that older adults are at higher risk of severe COVID-19 disease and COVID-19 mortality, providing a third dose to this population should be a top priority. Our data also highlight that understanding the waning of the immune response in other age groups is key for making evidence-based policies regarding booster vaccinations for the population at large.

## Introduction

The COVID-19 pandemic, which has been affecting global health for the past two years, is caused by severe acute respiratory syndrome coronavirus 2 (SARS-CoV-2). Age and frailty are among the strongest predictors of COVID-19 mortality.^1,2^ Due to immunosenescence, the primary vaccine response in those aged 65 years and older is associated with lower rates of protection compared to younger individuals.^3^ The BNT162b2 messenger RNA (mRNA) vaccine (Pfizer/BioNtech) induces generally lower antigen-binding IgG and virus-neutralising responses in individuals aged 65-85 years compared to those aged 18□55 years, when monitored two weeks post-vaccination,^4^ and that immunity wanes in all age groups.^5,6^ In July 2021, Israel became the first country in the world to authorise a third dose of BNT162b2 for adults aged 60 years and older.

We recently reported initial findings from a prospective cohort study that evaluated the anti-spike (anti-S) IgG antibody response before and after the third dose of the BNT162b2 vaccine in adults aged 60 years and older.^7^ We showed that the third vaccine dose was associated with significantly increased IgG titres, 10□19 days after that dose, with no major adverse events. The difference in median IgG titres before and after the third dose was >50-fold.^7^

The durability of response to the vaccine in adults aged 60 years and older is yet to be determined. Understanding the extent of waning immunity is critical for policy making, especially surrounding vaccination strategies. In this study, we evaluated the anti-S IgG antibody titres and neutralising antibodies three months after the third BNT162b2 dose in adults aged 60 years and older.

## Methods

### Trial oversight

The trial was conducted in accordance with the Declaration of Helsinki and was approved by the ethics committee of Rabin Medical Center (RMC). All participants provided written informed consent. The investigators were responsible for data collection and analysis.

### Study setting and participants

RMC is a tertiary hospital staffed by 7500 healthcare workers, including employees, students, and volunteers. Following the authorisation of a third dose in Israel on August 1, 2021, RMC offered this dose to the healthcare workers and their family members. Between August 4 and 12, study participants were recruited from those aged 60 years and older at the RMC vaccination centre. Exclusion criteria included prior SARS-CoV-2 infection (confirmed by PCR); steroidal treatment equivalent to 15 mg prednisone for the past 21 days or longer; active chemotherapy, immunotherapy, or biological treatments; active solid, hematologic malignancy, or both; and conditions affecting immunocompetence including liver cirrhosis, haemodialysis, solid organ transplant, bone marrow transplant, acquired immunodeficiency syndrome, inherent immune deficit such as congenital/acquired deficiencies of the complement system, asplenism or functional asplenism (e.g., Sickle cell disease).

### Sample collection

Blood samples were drawn from the study participants, before they received their third dose of the BNT162b2 vaccine (T0; August 4□12, 2021). Blood samples were also drawn 10-19 days (T1; August 16□24, 2021) and 74-103 days after the third vaccination (T2; November 3□15, 2021; except for two patients who came to their follow-up appointment on the wrong date, and for whom the blood sample was drawn on October 17, 2021).

In T2, a second blood sample was drawn from 60 participants who were randomly selected for neutralisation antibody analysis and sent to the Israel Institute for Biological Research (Ness Ziona, Israel) where the SARS-CoV-2 pseudovirus neutralisation assay was performed. The serum samples were stored in -80°C until the day of analysis.

### Assessments

Titres of anti-S IgG antibodies in the serum from the blood samples were determined at the RMC microbiological laboratory, using a chemiluminescent microparticle immunoassay, performed on the Abbott architect i2000sr platform, in accordance with the manufacturer’s package insert for SARS-CoV-2 IgG II Quant assay (Abbott Laboratories, Abbott Park, IL, USA; reference 6S60-22).^8^ The strength of the response (in relative light units [RLU]) was determined relative to IgG II calibrator/standard and reflects the quantity of IgG antibodies present. The assay is 98.1% sensitive 15 days or longer after the onset of COVID-19 symptoms or positive PCR test result and 99.6% specific.^9^ Seropositivity was defined as 50 arbitrary units (AU)/mL and higher.

Pseudovirus neutralising assay was performed using pseudoviruses expressing SARS-CoV-2 spike protein. Plasmids encoding a luciferase reporter (pGreenFire1, SystemBiosciences), lentivirus backbone (psPAX, Addgen), and S genes (Δ19 S-covid-pCMV3, a kind gift from Prof. Yossef Shaul, Weizmann Institute of Science, Rehovot, Israel) were co-transfected into HEK293T cells (ATCC CRL-3216). Forty-eight hours later, the medium was collected and virus aliquots were stored at -80°C for future use. One day before the pseudovirus neutralisation assay, hACE2-expressing HEK293 cells were plated in a white-wall 96-well plate (2×10^4^ cells per well). On the day of the assay, heat-inactivated sera were 2-fold serially diluted and mixed with pseudovirus, incubated for 1 hour at 37°C, and added to hACE2-expressing HEK293T cells. Twenty-four hours later, cells were lysed and luciferase activity (in RLU) was measured.^10,11^ Percent neutralisation was normalised using uninfected cells as 100% neutralisation, and cells infected with only pseudovirus as 0% neutralisation. IC50 titres were determined using a log (agonist) *vs* normalised-response (variable slope) nonlinear function using Prism software (GraphPad). Seropositivity was defined as a titre of 20 and higher.

The frailty of all participants was assessed at recruitment and confirmed at each timepoint thereafter via an interview using the 9-point clinical frailty scale (CFS).^12^ In addition, data were derived from the electronic medical records for all participants, including age, sex, vaccine doses and vaccination dates, and comorbidities.

Before the third vaccination and during both post-vaccination follow-up appointments, the study participants completed a questionnaire about adverse reactions post-vaccination and about whether they had a confirmed COVID-19 infection since the third dose/last follow-up and if so, their symptoms were documented.

### Statistical analysis

Participant characteristics were analysed using descriptive statistics. The difference in anti-S IgG values from T0 to T1, and from T1 to T2 was evaluated using Wilcoxon signed-rank test. Spearman correlation was used to assess the correlation between the anti-S IgG antibody values and neutralising antibodies titres. Univariate and multivariable analyses were performed by fitting a generalised linear model on the log of anti-S IgG antibody values at T2 and included age and days from the first vaccination as continuous variables, and sex, comorbidities (dyslipidaemia, hypertension, obesity, diabetes, and ischemic heart disease), and CFS as categorical variables.

For all analyses, IgG values above 80000 AU/mL were considered as 80000 AU/mL. A p-value of less than 0·05 was considered significant. All tests were two-sided. Statistical analysis was performed using R, version 4·0·2.^13^

## Results

Overall, 130 consecutive individuals aged 60 years and older were approached at the RMC vaccination centre, of whom one did not meet the eligibility criteria due to active malignancy and 28 refused participation. IgG levels at T0 were determined for 101 participants (78%). A total of four participants (3%) were lost to follow-up. Thus, the final cohort included 97 participants (figure 1). The median age was 70 years (IQR, 66□74), and 61% were women. The most common comorbidity was dyslipidaemia (61%) followed by hypertension (49%). The frailty of the majority of participants (58%) was characterised as “well” (CFS level 2) (table 1).

**Table 1.** Baseline demographics and cohort characteristics before and up to three months after the third BNT162b2 dose.

| Characteristic | N=97 |
| --- | --- |
| **Age** |  |
| Median (IQR), years | 70.0 (66–74) |
| **Sex, man, n (%)** | 38 (39) |
| **Comorbidities, n (%)** |  |
| Dyslipidaemia | 59 (61) |
| Hypertension | 48 (49) |
| Obesity | 26 (27) |
| Diabetes | 19 (20) |
| Ischemic heart disease | 17 (18) |
| Congestive heart failure | 1 (1) |
| **Clinical frailty scale, n (%)** |  |
| Very fit (level 1) | 28 (29) |
| Well (level 2) | 56 (58) |
| Managing well (level 3) | 8 (8) |
| Vulnerable (level 4) | 4 (4) |
| Mildly frail (level 5) | 1 (1) |
| **Analysis before the third dose (T0)** |  |
| Median (IQR) days after the first dose | 221 (218–225) |
| Median (IQR) IgG titres, AU/mL | 440 (294–923) |
| **Analysis 10-19 days after the third dose (T1)** |  |
| Median (IQR) days after the first dose | 236 (232–240) |
| Median (IQR) days after the third dose | 14 (14–17) |
| Median (IQR) IgG titres, AU/mL | 25429 (14203–36114) |
| **Analysis 74-103 days after the third dose (T2)** |  |
| Median (IQR) days after the first dose | 316 (312–320) |
| Median (IQR) days after the third dose | 94 (92–97) |
| Median (IQR) IgG titres, AU/mL | 8306 (4595–14701) |
| Median (IQR) neutralising antibody titre | 1,294 (848–2,072) |

AU=arbitrary units, IQR=interquartile range.

**Figure 1.**
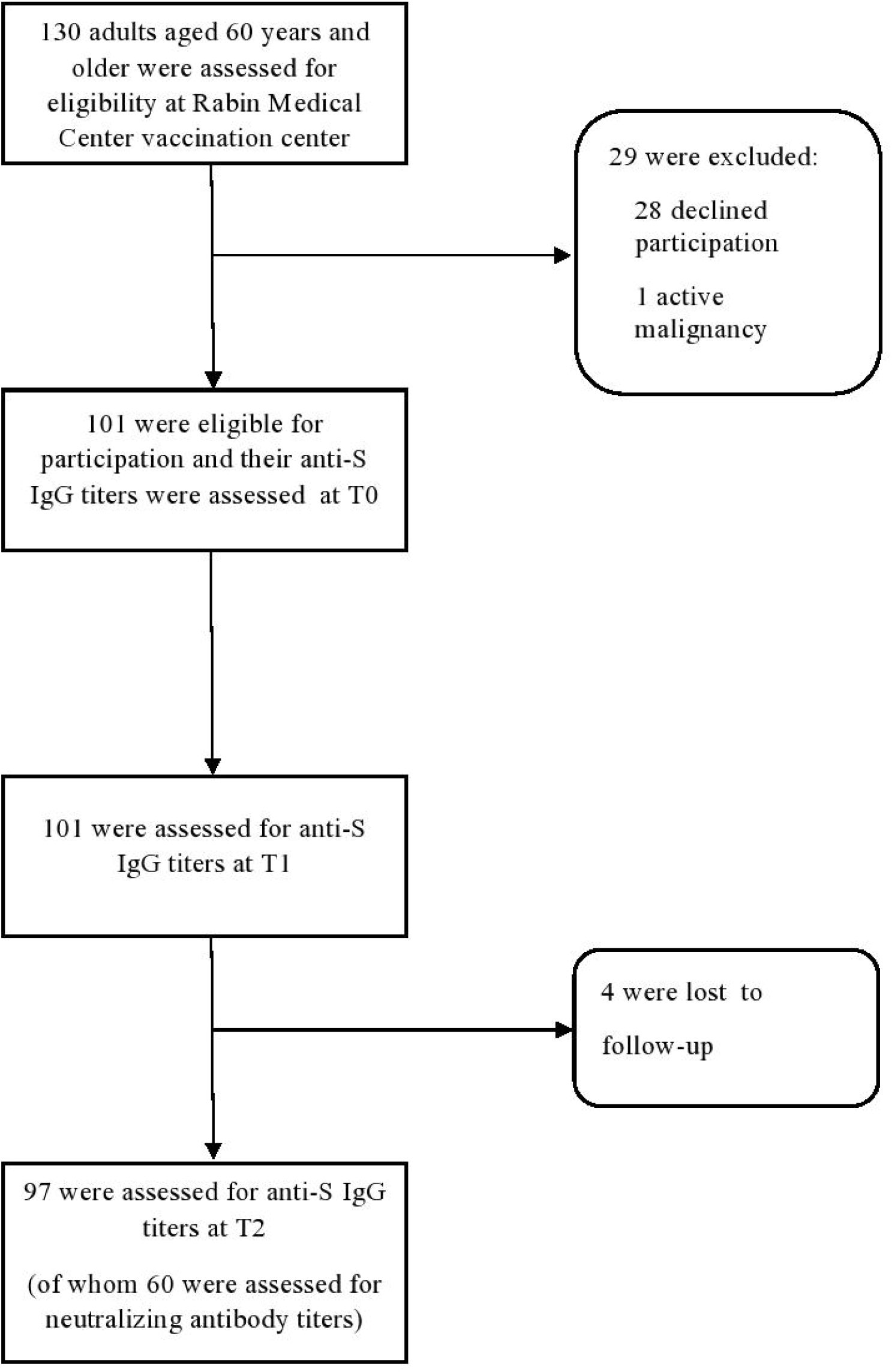
Study flow chart.

IgG titres, which increased significantly from before the third dose (T0) to a median of 14 days (IQR, 14□17) after the third dose (T1) (a median of 440 AU/mL [IQR, 294□923] *vs* 25,429 [14203□36114] AU/mL; p<0·001), decreased significantly approximately three months after the third dose (T2; a median of 94 days [IQR, 92□97] after the third dose), but all participants remained seropositive. At the T2 time point, the median IgG titre was 8306 AU/mL (IQR, 4595□14701) (p<0·001 *vs* T1) (Table 1, Figure 2).

**Figure 2.**
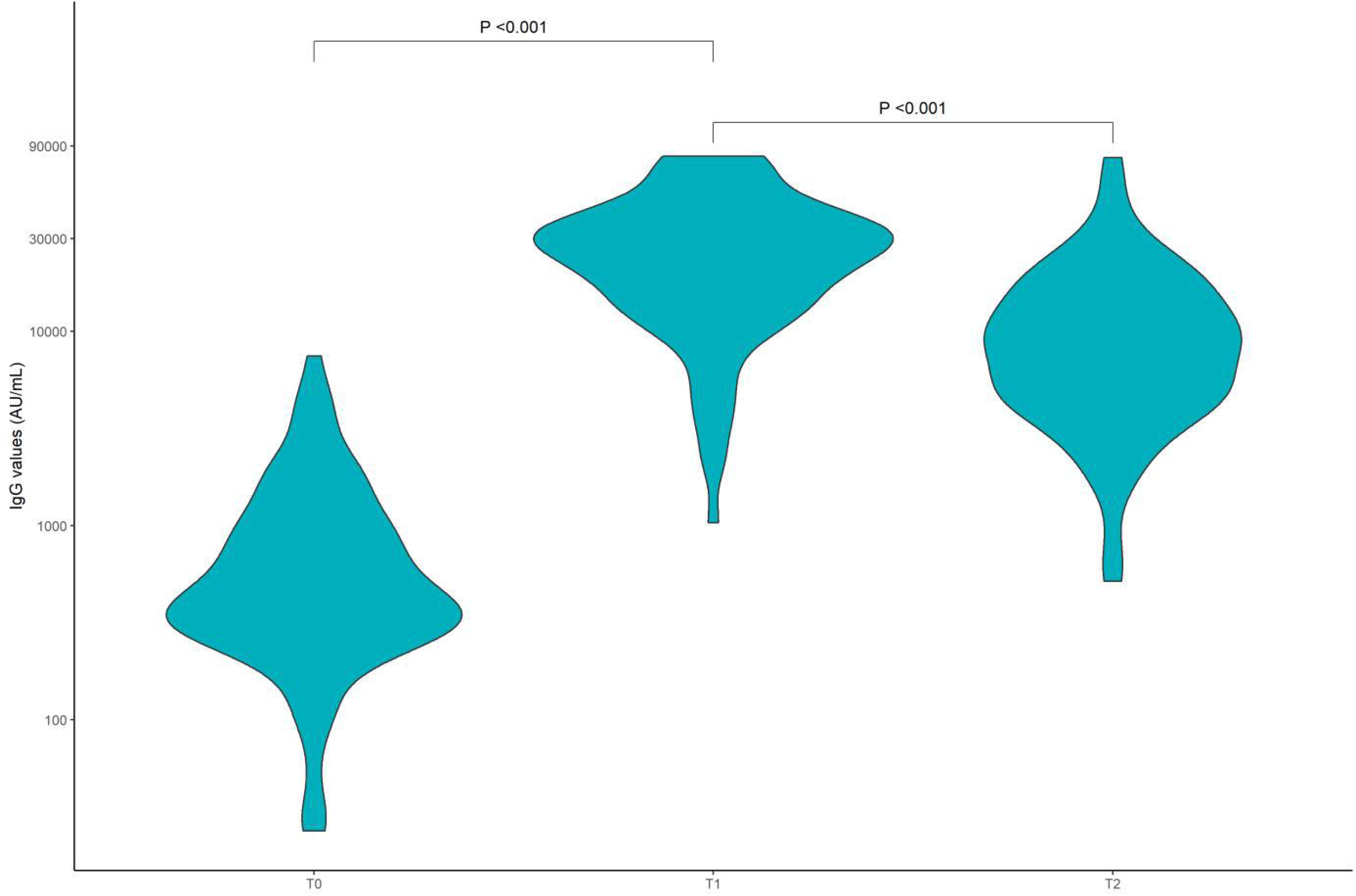
Anti-S IgG antibody titres over Time. Anti-S IgG titres were measured immediately before the third BNT162b2 dose (T0), a median of 14 days (IQR, 14-17) after that dose (T1) and a median of 94 (IQR, 92-97) after the third dose (T2).

In univariate and multivariable analyses, the only variable significantly associated with lower IgG levels at T2 was the number of days from the first vaccine dose (table 2).

**Table 2.** Univariate and multivariable analysis of log IgG values.

| Characteristic | Univariate analysis |  | Multivariable analysis |  |
| --- | --- | --- | --- | --- |
| | $\beta$ (95% CI) | p-value | $\beta$ (95% CI) | p-value |
| Age | -0.01 (-0.05 to 0.02) | 0.53 | 0.00 | 0.97 |
| Sex |  |  |  |  |
| Women | NA |  | NA |  |
| Men | -0.19 | 0.31 | -0.27 | 0.26 |
| Days from first vaccination | -0.04 (-0.07 to -0.01) | 0.009 | -0.04 (-0.07 to -0.01) | 0.004 |
| Comorbidities |  |  |  |  |
| Diabetes | -0.16 (-0.62 to 0.30) | 0.48 | -0.02 (-0.51 to 0.48) | 0.95 |
| Dyslipidaemia | -0.12 (-0.49 to 0.26) | 0.54 | -0.06 (-0.47 to 0.34) | 0.77 |
| Ischemic heart disease | -0.12 (-0.60 to 0.36) | 0.63 | 0.14 (-0.45 to 0.72) | 0.65 |
| Hypertension | -0.24 (-0.61 to 0.12) | 0.19 | -0.20 (-0.65 to 0.24) | 0.37 |
| Obesity | 0.05 (-0.36 to 0.46) | 0.81 | 0.24 (-0.18 to 0.67) | 0.26 |
| Clinical Frailty Scale |  |  |  |  |
| Very fit | NA |  | NA |  |
| Well | -0.19 (-0.60 to 0.22) | 0.40 | -0.06 (-0.53 to 0.42) | 0.81 |
| Managing well | -0.45 (-1.2 to 0.27) | 0.20 | -0.31 (-1.0 to 0.43) | 0.41 |
| Vulnerable | -1.0 (-1.9 to -0.04) | 0.045 | -1.0 (-2.0 to -0.04) | 0.044 |
| Mildly frail | 0.43 (-1.4 to 2.2) | 0.6 | 0.76 (-1.1 to 2.7) | 0.44 |

CI=confidence interval, NA=not applicable.

All participants for whom neutralising antibody levels were assessed (n=60) were positive for these antibodies. The median value of neutralising antibody titre was 1294 antibody titre (IQR, 848□2072). Evaluating the correlation between the anti-S IgG titres and neutralising antibody titres in these participants at T2 demonstrated a positive linear correlation (R=0·6, p<0·001) (figure 3).

**Figure 3.**
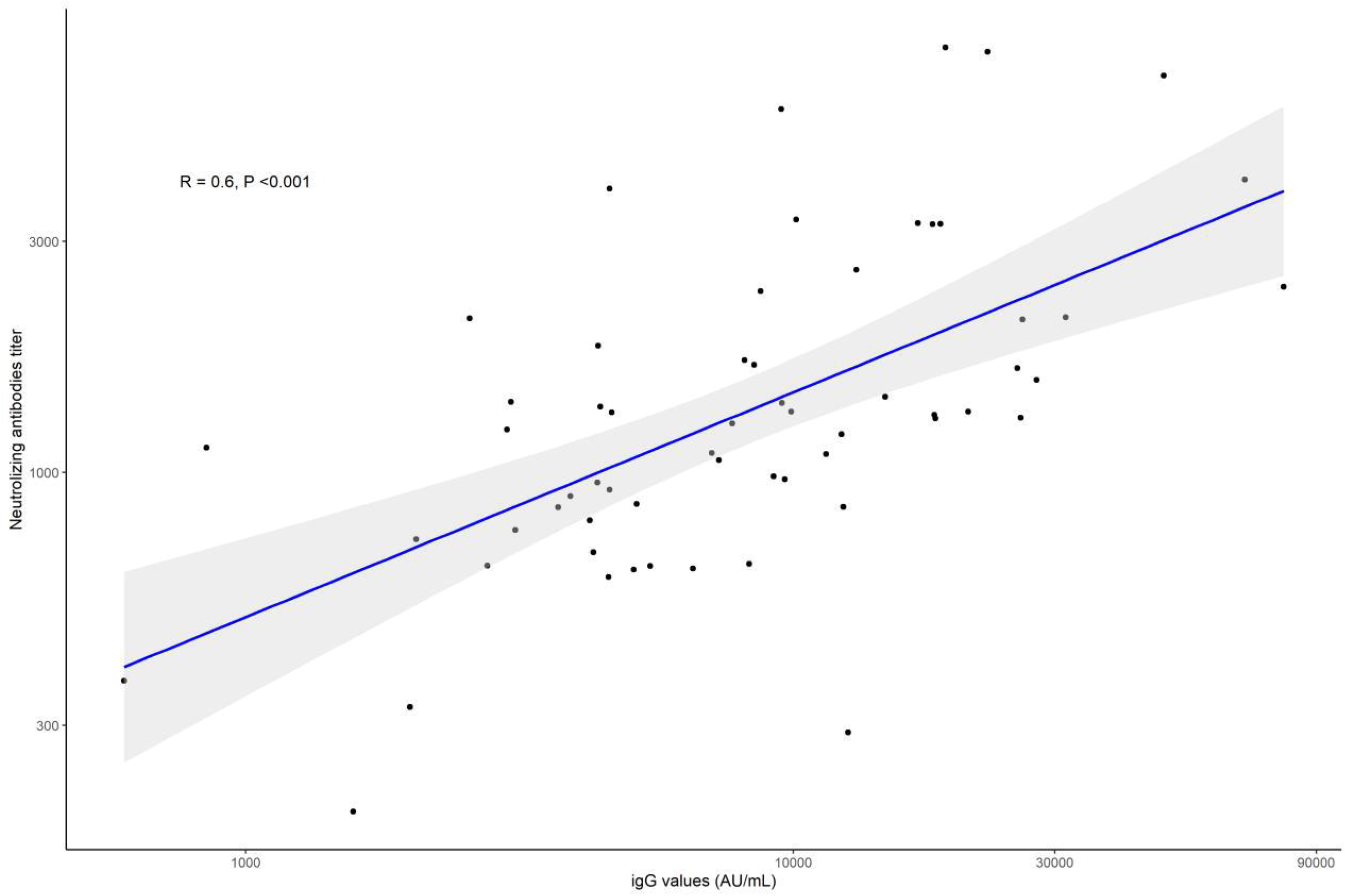
Neutralising antibody titres vs anti-S IgG titres at T2 (median of 94 [IQR, 92-97] after the third vaccine dose) in the 60 participants for whom both assessments were performed. Greyed areas represent 95% CI.

During the study period (median follow-up of 94 days [IQR, 92-97]), no major adverse events were reported and no participant had a COVID-19 infection. No change in frailty levels was observed in any of the participants throughout the study period.

## Discussion

This prospective cohort study demonstrated that anti-S IgG antibody levels increased significantly from before the third BNT162b2 dose to approximately two weeks after it (medians of 440 *vs* 25,429 AU/mL). However, approximately three months after that dose, a significant decrease in anti-S IgG levels was observed (median of 8306 AU/mL), although all participants remained seropositive.

In patients after natural COVID-19 infection, anti-S IgG levels were shown to be sustained, or progressively but moderately decline, whereas anti-receptor-binding domain of the spike protein (anti-RBD) IgG levels decline more commonly.^14^ One study showed that within 1·3 and 6·2 months of SARS-CoV-2 infection, titres of IgM and IgG antibodies against RBD decreased significantly while neutralising activity in the plasma decreased 5-fold in pseudotype virus assays ^15^ Neutralisation antibody dynamics was similar to that of anti-RBD antibodies in other studies as well.^16,17^ Overall, seropositivity rates remain high (88-90%) 6□8 months after natural infection.^18^

The clinical effectiveness of BNT162b2 against SARS-CoV-2 infection peaks in the first month after the second dose, declines gradually thereafter, and the decline accelerates after the fourth month.^19^ A recent study evaluated the long-term effectiveness of the BNT162b2 vaccine in participants of the phase 2–3 randomised trial and found a 4-fold decrease in its effectiveness between months 1–<2 vs 4–7 after the second dose (from 96% to 84%).^6^

The durability of protection after the third vaccination in healthy individuals at any age is still unknown. Understanding the extent of waning immunity is critical for public health policy making. To our knowledge, no serological follow-up beyond one month after the third dose has been published. The rapid waning of immunity,^5^ prompted investigation of the durability of the immune response after the third dose in order to assist decision-making regarding additional booster vaccinations.

Neutralising titres correlate with protection against infection, although the assays are complex and time-consuming.^20^ We found that all study participants were positive for neutralising antibodies in T2 with high titre levels and that the neutralising antibody levels were in correlation with anti-S IgG levels. Studies of other vaccines, such as measles, mumps and rubella, demonstrated a decrease of 5-10% in neutralising antibody levels per year.^21,22^ For COVID-19, the neutralisation level is highly predictive of immune protection. A recent study estimated that 50% protective neutralisation level equates to approximately 54 international units (IU)/mL (95% CI 30–96□IU/mL).^20^ Recent studies demonstrated that for those vaccinated at least five months earlier, a third BNT162b2 vaccine led to a rise in serum neutralisation titres by 5– to 7–fold,^23^ and was accompanied by 11.3–fold reduction in breakthrough infection rates.^24^ Comparing normalised neutralisation levels and vaccine efficacy demonstrated a strong non-linear relationship between mean neutralisation levels and the reported protection across different vaccines, including BNT162b2.^20^

It is well established that more than chronological age, the biological age (as reflected by frailty) is significantly associated with mortality.^25^ In COVID-19 patients aged 65 years and older, the CFS score was the strongest prognostic factor for mortality.^2^ In our cohort, the median age was 70 years (IQR 67–74), and the participants were ambulatory adults, mostly fit, as reflected by a median CFS of 2 (“well”) (IQR, “very fit” to “well”). Thus, the study population was young with respect to biological age. Investigating the waning immunity in a truly frail population is warranted.

No major side effects were observed in the three months after the third BNT162b2 vaccine, and none of the participants experienced a COVID-19 infection.

Lastly, it is impossible to discuss vaccine boosters without addressing the inherent ethical concerns. The current global environment manifests an imbalance in vaccine availability, with high-income countries delivering booster doses whereas the low-income countries are under-vaccinated with the first and second doses. Given the endemicity of this disease, waning vaccine protection, and the emergence of new variants, efforts are required to expand global vaccine access rapidly.^26^

Study limitations include small sample size, lack of cellular immunity testing and lack of neutralising antibody testing in the first two timepoints. Although the accumulating evidence suggests that IgG response and neutralising antibodies are correlates of disease protection,^27^ cellular immunity is also suggested to play an important role in protecting against SARS-CoV-2.^28^

In conclusion, in our cohort of 97 adults aged 60 years and older, three months after the third BNT162b2 vaccine, high levels of anti-spike and neutralising antibodies were found, but with significant waning of the immune response. Although further studies are needed to advance our understanding of waning immunity, the results suggest that a third vaccine dose for adults aged 60 years and older is effective and should be a top priority worldwide.

## Data Availability

All data produced in the present work are contained in the manuscript

## Contributors

NER, SMS, designed the trial and the study protocol. All authors contributed to data collection. AS performed the formal analysis. All authors had access to the data and SMS verified it. NER, SMS, and AB wrote the first draft of the manuscript. All authors reviewed it and contributed to the data interpretation and revisions. All authors had access to the data in the study, and SMS verified the data and the analysis. All authors reviewed and approved the submitted version and had final responsibility for the decision to submit the manuscript.

## Data sharing

This study is ongoing. The individual participant-level data that underlie the results reported in this article (text, tables, and figures) will be shared after de-identification, following the publication of the final endpoint of this study (6-month follow-up) upon request from the corresponding author.

**The study was not supported through external funding**.

